# Cohort profile: 10K - a large-scale prospective longitudinal study in Israel

**DOI:** 10.1101/2021.02.19.21251487

**Authors:** Smadar Shilo, Noam Bar, Ayya Keshet, Yeela Talmor-Barkan, Hagai Rossman, Anastasia Godneva, Yaron Aviv, Yochai Edlitz, Lee Reicher, Dmitry Kolobkov, Bat Chen Wolf, Maya Lotan-Pompan, Kohava Levi, Ori Cohen, Hila Saranga, Adina Weinberger, Eran Segal

## Abstract

The 10K is a large-scale prospective longitudinal cohort and biobank that was established in Israel. The primary aims of the study include development of prediction models for disease onset and progression and identification of novel molecular markers with a diagnostic, prognostic and therapeutic value. The recruitment was initiated in 2018 and is expected to complete in 2021. Between 28/01/2019 and 13/12/2020, 4,629 from the expected 10,000 participants were recruited (46%). Follow-up visits are scheduled every year for a total of 25 years. The cohort includes individuals between the age of 40 and 70 years old. Predefined medical conditions were determined as exclusions. Information collected at baseline includes medical history, lifestyle and nutritional habits, vital signs, anthropometrics, blood tests results, Electrocardiography (ECG), Ankle–brachial pressure index (ABI), liver US and Dual-energy X-ray absorptiometry (DXA) tests. Molecular profiling includes transcriptome, proteome, gut and oral microbiome, metabolome and immune system profiling. Continuous measurements include glucose levels using a continuous glucose monitoring (CGM) device for 2 weeks and sleep monitoring by a home sleep apnea test (HSAT) device for 3 nights. Blood and stool samples are collected and stored at −80 °C in a storage facility for future research. Linkage is being established with national disease registries.

## Why was the cohort set up?

The 10K is a prospective study conducted in Israel aimed at recruiting 10,000 individuals, who upon recruitment are aged 40-70 years old, and follow them longitudinally for 25 years. The primary goals are to study the variation observed across different individuals in disease susceptibility, clinical phenotypes, and therapeutic responses. We wish to analyse the complex interplay and relative contribution of genetic, epigenetic, microbiome and environmental factors on disease pathogenesis and progression and to evaluate how these effects are mediated with the goal of identifying novel molecular biomarkers with a diagnostic, prognostic and therapeutic value. We will develop risk prediction models for clinical outcomes based on data collected at baseline to evaluate the likelihood of an individual to develop clinical outcomes. Our initial focus will be on type 2 diabetes (T2D), cardiovascular disease (CVD) and obesity, as these conditions are highly prevalent, and earlier interventions such as lifestyle changes may prevent or delay their onset (1,2). In addition to the goals mentioned above, we believe that the rich dataset of deep and longitudinal phenotyping of participants collected in this cohort will enable the research of many additional scientific questions.

In recent years, there has been a rapid increase in the variety and capacity of large-scale epidemiological studies worldwide, often termed “biobanks”. As these biobanks are driven by different scientific goals, and as limited research resources often dictate tradeoffs between different axes of data collected (3), these studies vary in number of participants, length of follow-up and types of data collected (4). Compared to other biobanks, the 10K cohort has several unique aspects. First, while most of the cohorts collected to date have focused on genetics, we aim at obtaining a more comprehensive, person-specific, multi-omic molecular profile that includes genotyping, transcriptome, proteome, microbiome, metabolome and immune system profiling. By analysing different layers of data for each individual, collected longitudinally at multiple time points, we wish to reveal which omic layer is more perturbed and informative for each disease and to identify associations between molecular markers and health outcomes. Second, each participant in the cohort goes through a comprehensive profiling for two weeks at baseline, which includes continuous glucose monitoring coupled with self detailed logging of daily activities and continuous sleep monitoring for 3 nights. This data will allow us to track the dynamics and variation of these measures within the same individual and across different individuals in time at a high resolution, even in individuals who did not yet reach the thresholds defined as clinically significant. Third, the 10K cohort is one of the largest longitudinal studies established in Israel thus far. In analyzing the relative contribution of genetic and environmental influences, studying the Israeli population has an advantage as the Israeli population originates from several different ancestries who reside in a relatively small geographic region, and therefore share a relatively similar environment and habits (5). Finally, in parallel to the main 10K cohort, that will include relatively healthy individuals (see detailed exclusion criteria below), we also collect smaller cohorts of individuals with predefined medical conditions. These individuals go through the same process of data collection defined below, thereby allowing us to directly compare them to the healthy individuals at baseline and throughout the followup period. In these cohorts, we will also analyse factors associated with treatment modalities and disease prognosis. For example, at the moment we are collecting an additional cohort of individuals with cardiovascular morbidity. Similarly, we plan to collect cohorts of individuals with diabetes and oncology patients. These cohorts are planned separately, each with its own exclusion and inclusion criteria, and will not be further elaborated here.

## Who is included in the cohort?

The recruitment process relies primarily on self-assignment of volunteers who register to the trial website (https://www.project10k.org.il/en). Inclusion criteria included age range between 40-70 years old. Chosen similarly to the UKbiobank cohort (6), this age range will allow the investigation of common causes of morbidity and premature mortality, such as cardiovascular diseases and diabetes, with less comorbidity than observed at older ages.

Exclusion criteria were predefined by a team of expert physicians for the purpose of creating a relatively homogeneous study population, composed of individuals who were not yet diagnosed with outcomes of interest. Recent antibiotic usage and gastrointestinal morbidity were also defined as an exclusions due to their major influence on microbiome composition (7) (8). Participants are screened for the following exclusion criteria based on a designated questionnaire filled online:

- Pregnancy or currently undergoing fertility treatments
- More than 3 hospitalizations in the previous year
- Cardiovascular comorbidity such as myocardial infarction, congestive heart failure, valve dysfunction, cerebrovascular accident (CVA) or transient ischemic attack (TIA)
- Chronic neurologic or psychiatric comorbidity such as dementia, parkinson’s disease and amyotrophic lateral sclerosis (ALS)
- Chronic respiratory disease such as cystic fibrosis, interstitial lung diseases, ventilation need
- Chronic kidney disease such as renal tubular acidosis, urinary catheter usage or dialysis
- Chronic gastrointestinal disease such as inflammatory bowel disease (Crohn’s disease or Ulcerative Colitis) and liver cirrhosis
- Chronic metabolic diseases such as diabetes or adrenal insufficiency
- Antibiotic usage in the last 3 months
- Chronic antibiotic usage
- Weight loss > 5% in the recent year without deliberate diet
- Previous or active malignancy

Following the completion of the initial screening survey, individuals eligible for participation receive detailed information about study participation and an invitation to a comprehensive health assessment at the research site located at the Weizmann Institute of Science, Israel. Registration for the study at the 10K website (https://www.project10k.org.il/en) began on 28/10/2018. Between 28/01/2019 and 13/12/2020, 4,629 participants participated in the initiation meeting at the research site. Mean age was 51.50 ± 8.18 years old, median age was 50 years old (interquartile range 45-57). 326 (7%) participants have an additional family member participating in the study. Selected characteristics are shown in Table 1.

**Table 1:**
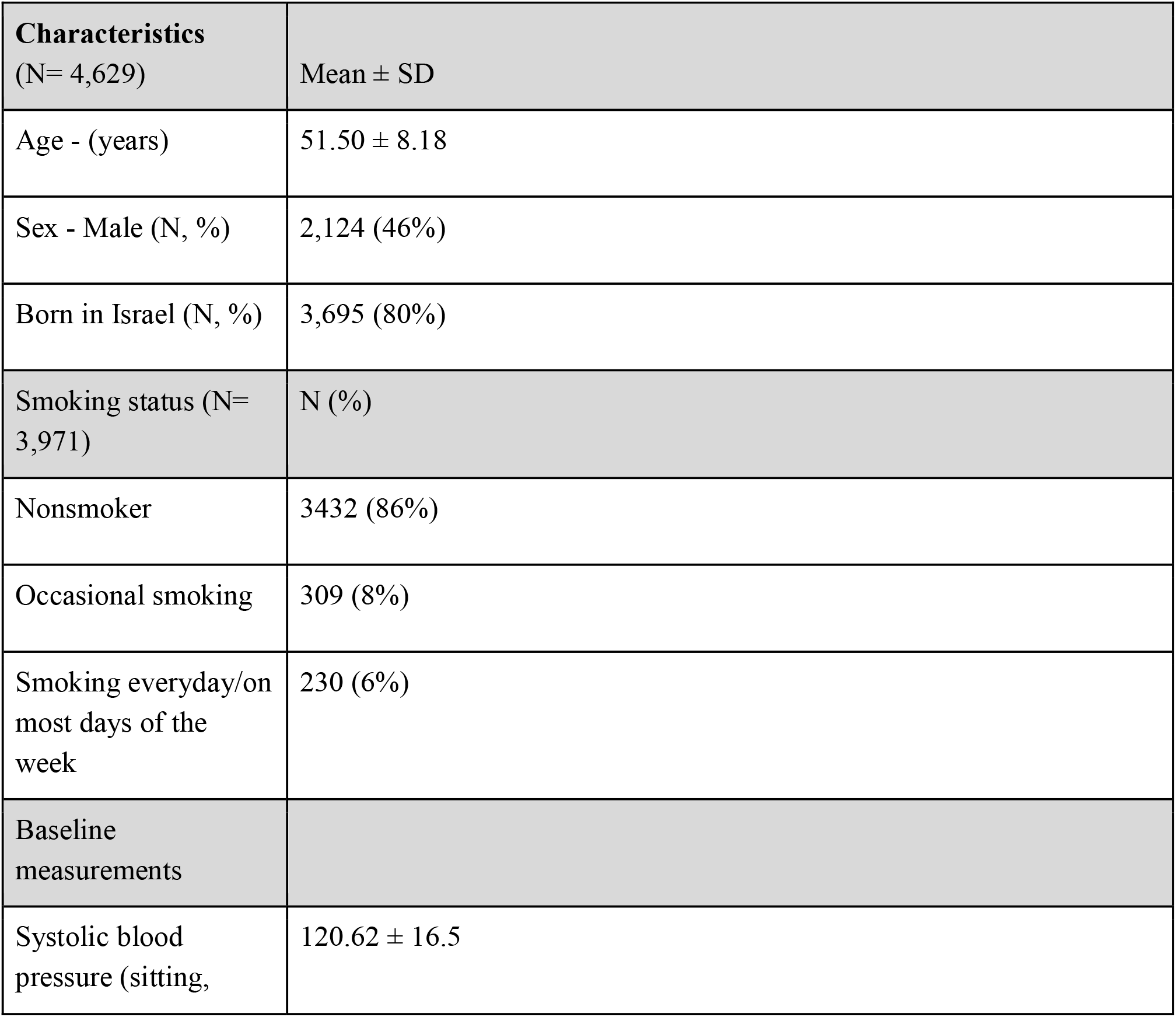
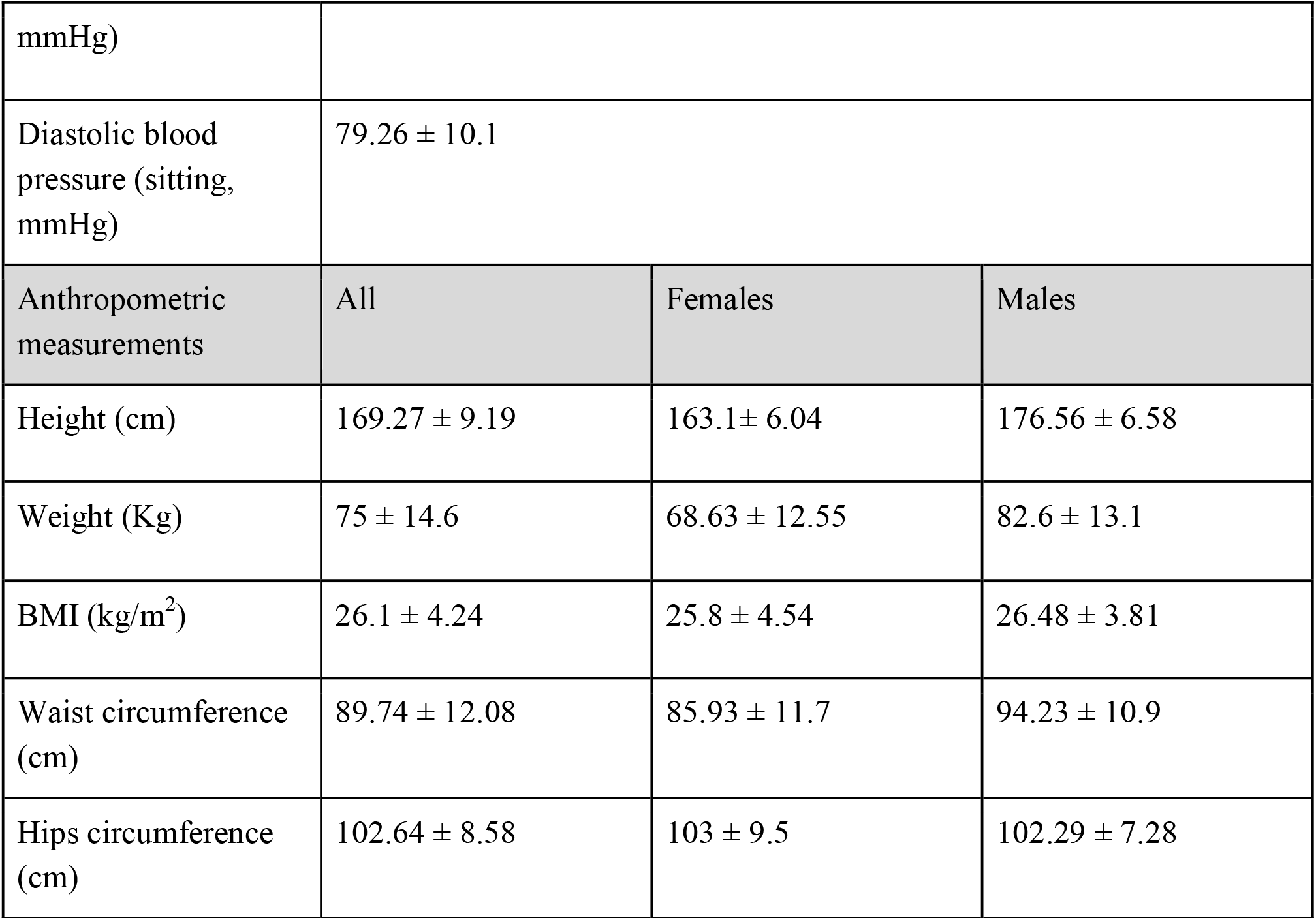
selected characteristics of the first 4,629 participants of the 10K study at baseline

| **Characteristics**<br>(N= 4,629) | Mean ± SD |
| --- | --- |
| Age - (years) | $51.50 \pm 8.18$ |
| Sex - Male (N, %) | 2,124 (46%) |
| Born in Israel (N, %) | 3,695 (80%) |
| Smoking status (N= 3,971) | N (%) |
| Nonsmoker | 3432 (86%) |
| Occasional smoking | 309 (8%) |
| Smoking everyday/on most days of the week | 230 (6%) |
| Baseline measurements |  |
| Systolic blood pressure (sitting, | $120.62 \pm 16.5$ |

|  |  |  |  |
| --- | --- | --- | --- |
| mmHg) |  |  |  |
| Diastolic blood pressure (sitting, mmHg) | 79.26 ± 10.1 | | |
| Anthropometric measurements | All | Females | Males |
| Height (cm) | 169.27 ± 9.19 | 163.1 ± 6.04 | 176.56 ± 6.58 |
| Weight (Kg) | 75 ± 14.6 | 68.63 ± 12.55 | 82.6 ± 13.1 |
| BMI (kg/m<sup>2</sup>) | 26.1 ± 4.24 | 25.8 ± 4.54 | 26.48 ± 3.81 |
| Waist circumference (cm) | 89.74 ± 12.08 | 85.93 ± 11.7 | 94.23 ± 10.9 |
| Hips circumference (cm) | 102.64 ± 8.58 | 103 ± 9.5 | 102.29 ± 7.28 |

## Ethical considerations

All participants sign an informed consent form upon arrival to the research site. All identifying details of the participants are removed prior to the computational analysis. The 10K cohort study is conducted according to the principles of the Declaration of Helsinki and was approved by the Institutional Review Board (IRB) of the Weizmann Institute of Science.

## What has been collected and measured?

Participants are invited to an initiation meeting in the research center. Measurements and samples obtained in the meeting are specified in Table 2. In addition, fundus imaging will be included for future participants. Thus far, nearly 37,000 samples (8 per participant) were collected. The baseline questionnaire includes questions on demographics, health status, lifestyle and psychosocial aspects (Table 3).

**Table 2.** Baseline measurements in the 10K study

| Measurement | Description and measurement technique |
| --- | --- |
| **Measurements obtained prior to the initiation meeting** |  |
| Blood tests | Participants are asked to provide recent and past results of the following blood tests prior to study initiation: Complete Blood Count (CBC), Hemoglobin A1C (HbA1c), thyroid stimulating hormone (TSH), C-Reactive Protein (CRP), creatinine, glucose, total cholesterol, High-density lipoprotein (HDL), low-density lipoproteins (LDL), triglycerides, albumin, protein, aspartate transaminase (AST), alanine transaminase (ALT), sodium, potassium, chloride, calcium, phosphor, vitamin B12 and vitamin D |
| **Measurements obtained in the initiation meeting at the research site** |  |
| Anthropometry | Height without shoes: Shekel stadiometer model H150-01-5<br>Weight : Shekel, model H150-01-5<br>Waist and hip circumference: measuring tape |
| Blood pressure | One measurement obtained after 5 minutes of chair-seated rest: OMRON blood pressure monitoring device |
| Orthostatic hypotension | One measurement obtained after 5 minutes laying down, followed by two measurements, one after 1 minute and another after 3 minutes standing up: OMRON blood pressure monitoring device |
| Blood tests | A total of 13 cc of blood is collected and stored in a robotic freezer for the analysis |
|  | of:<br>metabolomics<br>Transcriptome of peripheral blood mononuclear cells (PBMCs)<br>Serum proteomics<br>Antibody repertoire<br>Inflammation markers<br>Future assays |
| Electrocardiogram | 1 measurement with NORAV, 12 leads |
| Liver US | 1 measurement with Supersonic, Aixplorer MACH |
| Ankle–brachial pressure index (ABI) | 1 measurement with Viasonix - FALCON |
| US- doppler carotids for Intima Media Thickness (IMT) | 2 measurements, one for each side, with Supersonic- Aixplorer MACH |
| Pulse Wave velocity (PWV) | 1 measurement with Viasonix - FALCON |
| Bone density scan (DXA scan)<br>Total body composition scan | 1 measurement with GE- Lunar Prodigy Advance |
| Oral microbiome | 1 sample collected with a buccal swab.<br>Processing and analysis done by Gencove using low pass sequencing |
| Human genetics | 1 sample collected with a buccal swab.<br>Processing and analysis done by Gencove using low pass sequencing |
| Voice recording | 1 voice recording for a duration of 30 seconds conducted in the 10K research center in a designated room |
| Hand grip test | 2 measurements, one for each hand, by Jamar Hydraulic Hand Dynamometer |
| **Measurements obtained after the initiation meeting** |  |
| Gut microbiome | 1 stool sample is self collected by participants with DNA Genotek OM-200 kit |
| Continuous glucose measurements | Connection to FreeStyle Libre Pro Flash continuous glucose monitoring (FSL-CGM) system for two weeks |
| Continuous real-time, food intake, sleep times, and physical activity logging | Activity logging for two weeks by a proprietary smartphone App (available at https://play.google.com/store/apps/details?id=il.org.project10k.app) |
| Physical activity monitoring | Performed by health and activity tracking application installed on the mobile phone |
| Continuous sleep monitoring | Performed by a home sleep apnea test (HSAT) WatchPAT™ by Itamar Medical (9) for 3 nights |

**Table 3.** Overview of the content of the baseline questionnaires

| Topic | Dimensions (examples) |
| --- | --- |
| General information | Demographics, family composition, income, occupation, educational level, information regarding family relation to other participants |
| Health | Medical history, medication use, family medical history |
| Lifestyle and environment | Nutritional habits, smoking, sleeping habits, physical activity |
| Psychosocial parameters | Stress, quality of life, well-being |

## How long is the follow-up?

Participants will be followed up for 25 years. A follow-up questionnaire is distributed to all participants every year. A followup meeting at the 10K research site, in which repeated tests will be obtained will take place every two years. These meetings will include comprehensive follow-up surveys, repeated measurements and additional biobanking of samples. An overview of the study timeline is presented in Figure 2. Thus far, 2,593 participants have completed the follow up of the first year.

**Figure 1:**
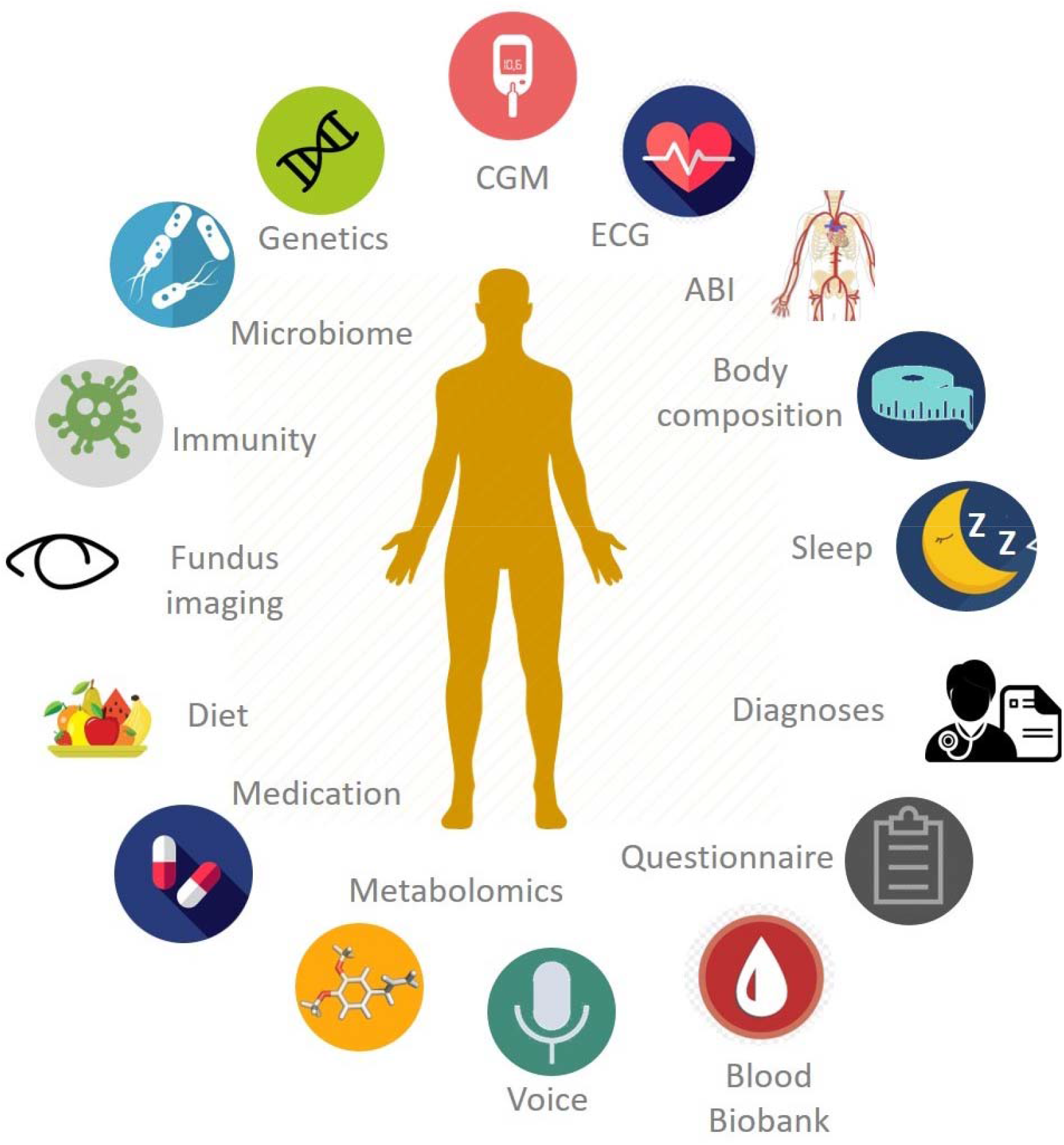
Data collected or analysed in the 10K cohort

**Figure 2:**
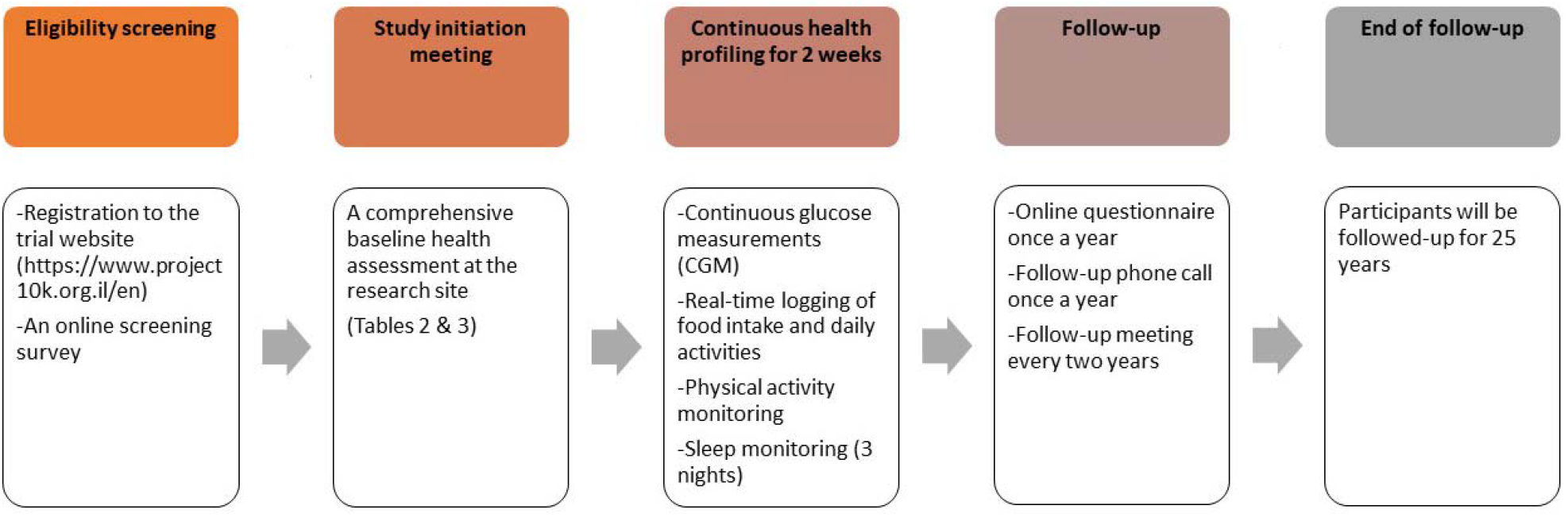
Timeline of the 10K cohort

## Key findings

Prior to establishing the 10K project, we have collected data on 900 healthy individuals who were phenotyped for medical background, physical measures, lifestyle habits, genetics, oral and gut microbiome and serum metabolites and were connected to a CGM device in parallel to logging meals in a proprietary smartphone App for two weeks. This preliminary cohort enabled us to inquire into several scientific questions, such as the variability in postprandial glycemic responses among healthy individuals (10), the relative contribution of genetics versus environmental factors in shaping gut microbiome composition (5), the associations between microbial genomic structural variants and clinical phenotyping (11) and the potential determinants of blood metabolites (12). In the 10K cohort, we are expanding the number of participants as well as adding new types of assays to create a deeper phenotypic profile for each participant along with longitudinal follow-up. We therefore expect that data collected here will enable the pursuit of many additional scientific endeavors.

## What are the main strengths?

The strengths of this study include its prospective design and long-term follow-up; the relatively large sample size, which will provide sufficient statistical power to study prevalent health outcomes, reliably and reassessment of the health status of participants and repeated measurements throughout the followup period. An additional strength is the deep phenotyping of each participant, which includes many layers of molecular data as well as continuous measurements which can facilitate a wide range of scientific questions. For example, despite many studies conducted on the human microbiome and its role in numerous diseases (13), very few microbiome-based markers that are predictive of disease onset and progression were found to date, making it a promising yet mostly unexplored area of research.

## What are the main weaknesses?

The limitations of the study include its observational nature, which limits the ability to infer causation. We will try to address this challenge by using different strategies such as Mendelian randomization (14) and employing causal inference methods (15,16). Biomarkers identified as having a potential therapeutic value will need to be tested in randomized clinical trials to establish causality and be assessed for their safety and efficacy. However, we hypothesize that testing omic targets already shown to be associated with clinical outcomes in humans will make them more likely to succeed in clinical trials, as was previously shown in drug targets identified by genetic evidence (17). Another limitation in this cohort, as well as others (18), is the possibility of selection bias, that may influence the ability to generalise associations of exposure with disease (19). The size of the cohort is also limited compared to other nationwide cohorts, but is still relatively large for a deeply phenotyping cohort that includes all of the above physiological and molecular assays. Finally, although linkage is being established with national disease registries, linkage to the participants’ Electronic Health Record (EHR) has not been established to date, and therefore information on clinical outcomes is obtained through self-reported surveys and phone inquiries by the research staff.

## Can I get hold of the data? Where can I find out more?

Access to the 10K Cohort is currently not available online. Potential collaborators are encouraged to contact the Principal Investigator by e-mail for further information.

## Data Availability

Access to the 10K Cohort is currently not available online. Potential collaborators are encouraged to contact the Principal Investigator by email for further information.

## Acknowledgements

The authors wish to acknowledge all participants of the cohort and the members of the Segal lab for fruitful discussions.

